# Multi-branch convolutional neural network using intracranial EEG high frequency oscillation features for predicting post-surgical seizure outcomes

**DOI:** 10.1101/2025.10.05.25337367

**Authors:** Madhumathi Devaraj, Yihe Chen, Shuang Wang, Michael R. Sperling, Noa Herz, Chengyuan Wu, Richard Staba, Jerome Engel, Itzhak Fried, Charles Mikell, Sima Mofakham, Petar Djuric, Shennan Weiss

## Abstract

Pathological high-frequency oscillations (HFOs 80-600 Hz) in intracranial EEG distinguish epileptogenic cortex. However, it is uncertain whether utilizing HFO measures for surgical planning improve epilepsy surgery seizure outcomes and minimize morbidity. The clinical gold standard for planning an epilepsy surgery involves consensus between epileptologists, radiologists, and neurosurgeons based on multimodality findings, and particularly the location of the seizure onset zone. We asked whether seizure freedom following epilepsy surgery could be accurately predicted using machine learning that uses measures of HFO features relative to the boundaries of a surgical resection or laser ablation. We detected and quantified HFOs from depth EEG contacts during 30-200 minutes of non-rapid eye movement sleep from 78 pre-surgical patients from three institutions. We trained a three-branch convolutional neural network (CNN) using 3 neuroanatomic features and 37 HFO derived features. The first and second CNN branches computed within and between patient differences, respectively, and the third branch contains the resected contacts that also influenced branches 1 and 2. We found that this HFO CNN labeled the seizure free patients with 92% accuracy using 5-fold cross-validation. These results suggest that a resection planned with the clinical gold standard can be prospectively evaluated by a HFO CNN approach to test whether the resection boundaries will achieve a seizure free outcome. Future work will explore utilizing the HFO CNN approach for counterfactual virtual resections constrained by a utility function to minimize morbidity.

## Introduction

Epilepsy is a serious and common neurological disorder characterized by spontaneous seizures and increased risk of sudden death, especially for one-third of the individuals with chronic seizures that cannot be controlled despite optimal drug therapy. Many of these medically refractory patients are candidates for epilepsy surgery. One type of epilepsy surgery involves the resection or thermal ablation of brain tissue thought to be necessary and sufficient for seizure generation.

The gold standard for defining the boundaries of surgical resection is determined by the collective judgment of a multidisciplinary team. This consensus is informed by an integrated analysis of multimodal data particularly localizing the electroclinically-defined seizure onset zone (SOZ)[1]. Important biomarkers for predicting post-operative seizure outcomes in patients undergoing epilepsy surgery include simplified clinical metadata[2], [3], radiographic data[4], and more detailed neurophysiological data alone[5], [6]. In this study machine learning was used to predict post-operative seizure outcome. The features used for the machine learning framework are derived from specific neurophysiological biomarkers called high-frequency oscillations (HFOs: 80-600 Hz). HFOs are brief bursts (8-200 msec) in spectral energy subcategorized as ripples (80-200 Hz) and fast ripples (200-600 Hz). Ripples and fast ripples are most often identified in the intracranial EEG (iEEG) in patients undergoing invasive monitoring for planning epilepsy surgery during non-rapid-eye-movement (NREM) sleep[7]. A substantial body of research spanning the last 25 years has robustly established a significantly higher rate of High-Frequency Oscillations (HFOs) within the seizure-onset zone (SOZ)[8], [9], [10], [11], [12], [13], [14], [15]. However, surgical resection of the SOZ fails to produce seizure freedom in 35-50% of patients, and this failure rate is even greater for those with MRI-negative (non-lesional) epilepsy[16], [17], [18].

A significant conceptual advance in epilepsy surgery was the introduction of the epileptogenic zone (EZ). The EZ is defined as the hypothetical brain region necessary and sufficient for seizure generation[19]. Consequently, a surgery that renders a patient seizure-free is presumed to have completely resected the EZ[19], [20]. High-Frequency Oscillations (HFOs)—including ripples and fast ripples, particularly when superimposed on epileptiform spikes[15], [21] —have been proposed as biomarkers for the EZ[5], [6], [22]. Supporting this, studies consistently show that patients achieving post-operative seizure freedom have a significantly higher resection ratio of HFO-generating tissue compared to those with recurrent seizures [5], [6], [22]. These findings indicate that the EZ, identified by HFOs, is not always spatially congruent with the SOZ and could extend beyond it. Therefore, targeting this HFO-generating network is critical for a successful surgical outcome.

In this study, using iEEG recordings during NREM sleep from a cohort of 78 patients implanted with depth electrodes, we asked if a multi-branch convolutional neural network (CNN) utilizing features including the HFO rates and other HFO derived features measured per depth electrode contact could predict which patients would achieve post-operative seizure freedom. These measures were examined both in a neuroanatomical context and with respect to the margins of the resection or ablation.

## Methods

### Patients

Recordings selected from 7 patients who underwent intracranial monitoring with depth electrodes between 2014 and 2018 at the University of California Los Angeles (UCLA), from 15 patients at the Thomas Jefferson University (TJU) in 2016–18, and from 56 patients at Zhejiang University (ZJU) Hospital in China for the purpose of localization of the SOZ. Data collection was planned before the study was conceptualized. These 78 patients subsequently underwent resections or ablations. Inclusion criteria for this study included: 1) pre-surgical MRI for guided stereotactic electrode implantation; 2) a post-implant CT scan to localize the electrode contacts; 3) SEEG recordings during non-REM sleep at a 2-kHz sampling rate, and 4) a post-operative MRI. All patients gave verbal and written informed consent prior to participating in this research, which was approved by the UCLA, TJU, and ZJU institutional review boards. Eligible patients were found through queries of pre-existing clinical databases. The methods in this paper adhered to, and were in accord with, the relevant guidelines and regulations of the institutional review boards. Stony Brook University deemed this study IRB exempt and approved the data sharing agreements.

### Neuroimaging analysis

For each patient in the UCLA and TJU cohorts, pre-implantation and post-resection T1-weighted magnetic resonance images (MRIs) were acquired and processed using a custom pipeline (https://github.com/pennmem/neurorad_pipeline). Post-implantation computed tomography (CT) scans, obtained following the placement of stereoelectroencephalography (SEEG) depth electrodes and co-registered with the anatomical MRIs. This co-registration and subsequent normalization to a standard template were performed using Advanced Neuroimaging Tools (ANTs) [23] under the supervision of a neuroradiologist. The spatial coordinates of each electrode contact were then localized within the Montreal Neurological Institute (MNI) normalized space and anatomically labeled according to the Desikan-Killiany atlas [24]. Following co-registration, electrode contacts located within the resection cavity were identified manually using itk-SNAP. For the ZJU patients, the normalized MNI coordinates were reconstructed using Brainstorm (https://neuroimage.usc.edu/brainstorm/) using the maff-8 function to coregistrater the MRI brain and post-implantation CT and identify the resected contacts.

### EEG recordings and HFO detection

For each patient, clinical intracranial EEG (iEEG) was acquired from 8 to 16 depth electrodes (each with 7–15 contacts) using a 256-channel Nihon-Kohden JE-120 monitoring system (Nihon-Kohden America, Foothill Ranch, CA, USA) at UCLA, TJU, and ZJU. Data were recorded with a bandwidth of 0.1–600 Hz and a sampling rate of 2000 Hz. Implantation strategies varied by institution; a higher density of electrodes and contacts was typically used at TJU. The referencing scheme was also site-dependent: recordings at UCLA were referenced to a scalp electrode at position Fz, whereas TJU and ZJU recordings were referenced to an electrode contact in the white matter. For analysis, a 60–200-minute segment of iEEG data, recorded one to two days post-implantation, was selected from a period of non-REM (NREM) sleep, characterized by high-amplitude, delta-frequency slow waves. High-frequency oscillations (HFOs), categorized by frequency as ripples (80– 200 Hz) or fast ripples (200–600 Hz) superimposed on the iEEG background (Ripple on Oscillation [RonO], Fast Ripple on Oscillation [fRonO]) or on spikes (Ripple on Spike [RonS], Fast Ripple on Spike [fRonS]), as well as sharp-spikes generated by Gibb’s phenomenon[25], were automatically detected from the NREM sleep data using previously validated methods (https://github.com/shenweiss)[15], [26], [27], [28], [29] implemented in MATLAB[30]. Specifically, HFOs were identified and quantified via the topographical analysis of the wavelet convolution (Fig. 1)[26]. Following automated detection, all putative events were visually reviewed in Natus™ Brain Quick™ (Venice, Italy). This semiautomated analysis examined the annotated recordings in 10-minute blocks. Blocks contaminated with arousals associated with muscle artifact and decreased slow waves were excluded from further analysis. An additional inclusion criterion for the study was at least three uncontaminated 10-minute blocks.

**Figure 1:**
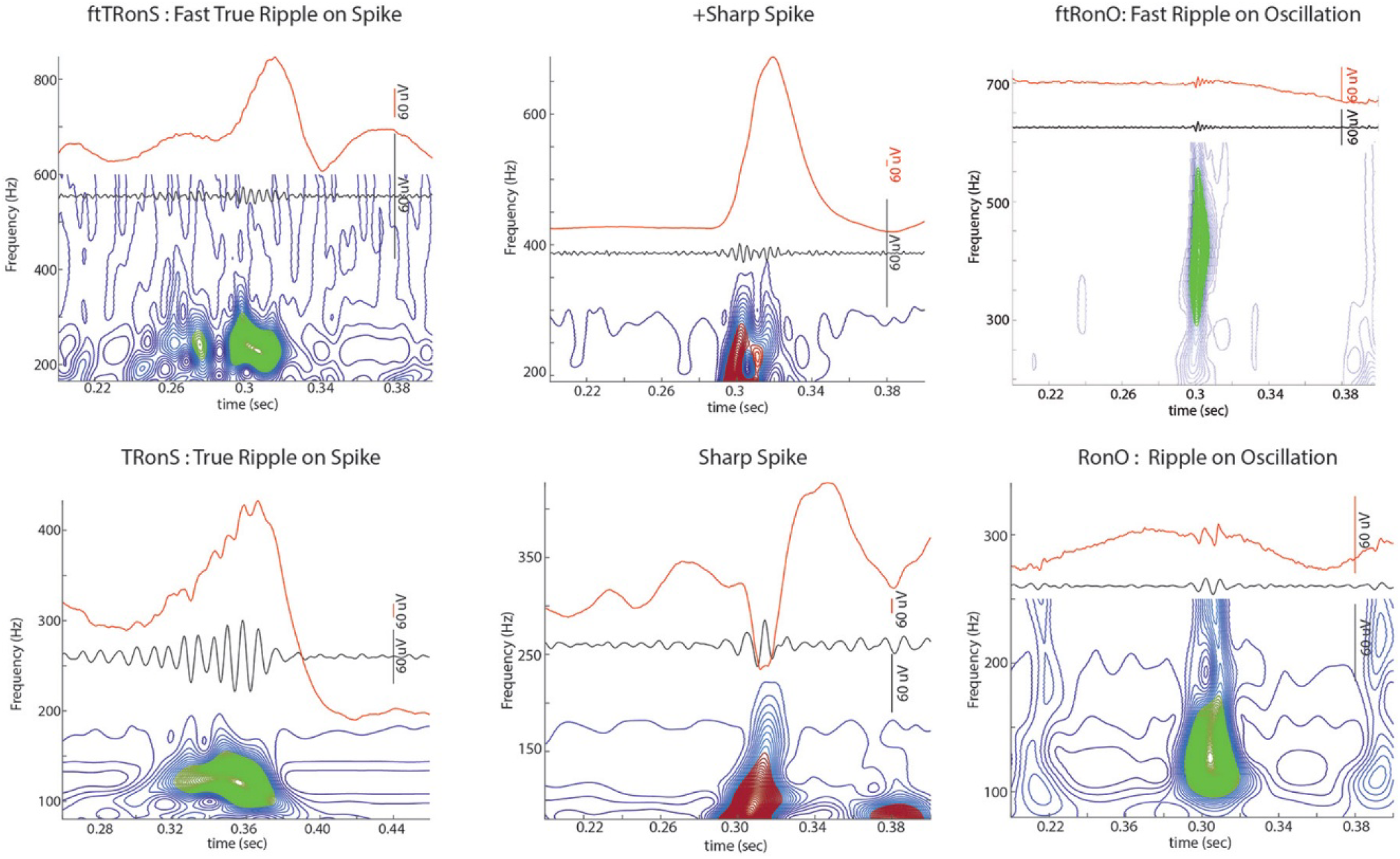
HFO subtypes quantified by the topographical analysis of the wavelet convolution. Creative Commons website: https://creativecommons.org/licenses/by/4.0/.

### Convolutional Neural Network Model Architecture

Based on clinical data obtained at least one year following epilepsy surgery (resection or ablation), using Engel outcome criteria, each patient was designated a binary outcome (0 = seizure free, 1 = not-seziure free). For each patient electrode contact label features included: 1) spatial coordinates in normalized MNI space (x, y, z); 2) rates of RonO, RonS, fRonO, fRonS, sharply contoured spikes (events/min) [12], [13], [21], [31], [32]; 3) HFO statistics (mean, stdev, and value at a z=3 threshold) related to the spectral content of these HFO events[14], [33] and the power of these HFO events (RonO, RonS, fRonO, fRonS)[34] 4) the rate of ripples superimposed on fast ripples [22]; 5) The rate wherein ripples [35] and fast ripples [36], [37] precede epileptiform discharges within 300 msec; 6) Ripple and fast ripple ratios during UP-DOWN vs. DOWN-UP transitions during slow and delta waves of sleep[27], [36], [38], [39], [40]; and 7) ripple and spike phase coupling for RonS events[14]. Items 2-7 compose the 37 HFO related features. Lastly, the electrode contacts were labeled by their resection or ablation status per electrode. Last (0 = unresected, 1=resected)

Preprocessing involved merging the seizure free outcome to the patient’s electrode contact level feature. A three-branch convolutional neural network (CNN) was designed in TensorFlow/Keras to integrate electrode-level features, aggregated statistics, and resection summaries. The model architecture included the following inputs: 1) Electrode input: shape (max_electrodes, n_features) for each patient; 2) Resection flag input: shape (max_electrodes, for each patient; 3) Mask input: shape (max_electrodes,) for padding; 4) Aggregated vector input: shape (agg_len,) of the 37 HFO related features across all patients for branch 2; and 5) Resected summary input: shape (3,) for each patient for branch 3. To calculate agg_lenn (i.e., #4) for branch 2 per-patient aggregation, for each patient, was performed by calculating statistics across the patient’s electrode level features. These per-patient aggregations included the mean and standard deviation of each feature across all the electrodes, mean of the features only in the resected electrodes, and counts of total electrodes, resected electrodes, and fraction of electrodes resected. These aggregated values summarize both the global patterns and surgical details at the patient level.

Values used by Branch 1 computed electrode contact-specific features while considering resection information from Branch 3. The values were standardized using StandardScaler in scikit-learn, across all contacts and within features, and. electrode features are concatenated with resection flags and passed through two 1D convolutional layers (kernel size=3, filters optimized using Keras Tuner). The resulting feature maps are transformed via dense layers to generate query and key vectors for attention. Attention weights are masked using the mask_input to ignore padded electrodes. A weighted sum over all electrodes produces a fixed-length patient-level representation capturing spatial and electrophysiological patterns.

Values used by Branch 2 included the aggregated vector input: shape (agg_len,) across all patients. StandardScaler from scikit-learn was used to standardize these values across patients. Branch 2 encodes these patient-level statistical features aggregated across electrodes. Features are passed through a dense layer with ReLU activation, followed by dropout to prevent overfitting. This branch captures global patterns and complements the fine-grained electrode-level representation from Branch 1.

Branch 3 encodes surgical resection information using a compact input vector containing the total number of electrodes, the number of resected electrodes, and the fraction of electrodes resected. This branch processes the input through a dense layer with half the number of units (dense_units/2), followed by ReLU activation and dropout. The reduction in units reflects the smaller size and lower dimensionality of the resection summary compared to the aggregated features, preventing overparameterization while preserving meaningful representation.

In the final layers, after processing through the three separate branches—Branch 1 (electrode-level CNN with attention), Branch 2 (aggregated patient-level features), and Branch 3 (resection summary), their outputs are concatenated into a single feature vector. This concatenation allows the model to integrate spatial-temporal electrode information, global feature statistics, and surgical resection data into a unified representation. The combined vector is then passed through a fully connected (dense) layer with ReLU activation and dropout. This layer serves two purposes: it learns higher-level nonlinear interactions among the three feature types, and dropout regularizes the network to reduce overfitting. Finally, the network outputs a single value through a dense layer with a sigmoid activation function. This output represents the predicted probability of the patient not being seizure-free post-surgery. By thresholding this probability at 0.5, the model produces the binary classification outcome, completing the prediction of post-surgical seizure outcomes.

### Convolutional Neural Network Model Architecture: Hyperparameter Tuning

Hyperparameter optimization was performed using Keras Tuner (keras_tuner package) with the Hyperband algorithm. Each fold in the stratified cross-validation underwent an automated search to select optimal values for: 1) Conv1D filter count (16–64); 2: Dense units (32–128); 3) Dropout rate (0.2–0.5); and 4) Learning rate (1e-3, 3e-4, 1e-4). All hyperparameters were tuned automatically. Hyperband efficiently evaluates multiple configurations by allocating more resources to promising models and pruning underperforming ones. This ensures that the search identifies optimal hyperparameter settings for model performance, while minimizing computational time and resource usage.

### Convolutional Neural Network Model Architecture: Training Procedure

The model was trained using a stratified 5-fold cross-validation scheme based on patient IDs. Stratification ensures that the distribution of seizure outcomes is preserved across folds, while using patient IDs prevents data leakage across electrodes from the same patient. Training was performed with a batch size of 8 and up to 50 epochs per Hyperband tuner trial. Early stopping was disabled to allow the model to complete full epochs, ensuring reproducibility of results across repeated experiments. To maintain deterministic behavior, random seeds were fixed at 42 across NumPy, TensorFlow, and Python’s random module. Training leveraged a GPU environment with CUDA 11.6 and TensorFlow-GPU to accelerate computation. This setup enabled efficient training of the multi-branch CNN while ensuring consistent and reproducible outcomes.

### Evaluation Metrics

TensorFlow/Keras were used to generate a receiver operating characteristic (ROC) curve based on the probability of the seizure free and non-seizure free labels and to calculate the area under the ROC. At a threshold probability of 50% confusion matrices were generated, and the following values were computed: 1) Sensitivity = (TP+FN)/TP; 2) Specificity = (TN+FP)/TN; 3) positive predictive value (PPV)=(TP+FP)/TN; 4) negative predictive value NPV=(TN+FN)/TN; 4) Accuracy = (TP+TN)/(Total); 5) Precision = TP/(TP+FP); 6) Recall = TP/(TP+FN); 7) F1-score = 2×Precision×Recall/(Precision+Recall); and 8) Youden’s J=Sensitivity+Specificity−1

### Univariate analysis of the 37 HFO related features

The patients with post-operative seizure free outcomes and those with non-seizure free outcomes were split into two distinct cohorts. The mean and standard error of each of the 37 HFO related features were calculated within the resected and unresected contacts for each of the two cohorts. Also, for each of the two cohorts, the 37 HFO feature values were individually compared between resected and unresected contacts using a unpaired t-test corrected for multiple comparisons (n=74) using the Holms-Bonferroni method. A Cohen’s d value was also derived for each of these 72 comparisons.

## Results

iEEG data was inspected in discrete 10-minute blocks and blocks with artifact were excluded. We included 4.86 +/- 1.21 (mean +/- std. dev.) blocks for the 7 UCLA patients, 10.53 +/- 5.0 blocks for the 15 TJU patients, and 7.67 +/- 3.30 blocks for the 56 ZJU patients. The 7 UCLA patients had 58.86 +/- 8.8 total contacts, the 15 TJU patients had 146.4 +/- 29.37 contacts, and the ZJU patients had 113.25 +/- 25.54 contacts. With respect to the number of contacts with iEEG including HFO events, the 7 UCLA patients had 47.14 +/- 7.38 total contacts, the 15 TJU patients had 110.6 +/- 23.63 contacts, and the ZJU patients had 97.4 +/- 19.3 contacts. Among the total contacts, the number of resected contacts was 12.86 +/- 3.4 for the 7 UCLA patients, 23.27 +/- 19.25 for the 15 TJU patients, and 19.625 +/- 10.67 for the 56 ZJU patients.

Across all the patients and analyzed blocks we detected 7,229,958 ripples on the iEEG background (i.e., RonO), 440,941 ripples on epileptiform spikes (i.e., RonS), 163,947 sharp spikes without HFO, 277,590 fast ripples on the iEEG background (i.e., fRonO), 93,566 fast ripples on epileptiform spikes (i.e., fRonS), and 4609 very sharp spikes without HFO (Fig. 1). We distinguished patients rendered seizure free by surgery from those with recurrent post-operative seizures and within each cohort computed the mean and standard error of each HFO feature in resected contacts and unresected contacts and compared the raw values with a t-test to assess significance and a Cohen’s d to compute effect size (Supplementary Table 1 and 2).

We found that the three branch convolutional neural network (CNN) using intracranial EEG HFO features in a neuroanatomic framework that included the resected electrodes predicted post-operative seizures following surgery with an accuracy of 92.3%, precision of 96%, recall of 82.8%, F1 score of 89%, sensitivity of 83%, specificity of 98%, positive predictive value of 96%, negative predictive value of 91% and an area under the receiver operating curve of 90% (Fig 2). By generating confusion matrices, at the maximum of Youden’s J, we also compared the overall performance of the CNN across all patients and the patients stratified by institution (Fig 3).

**Figure 2:**
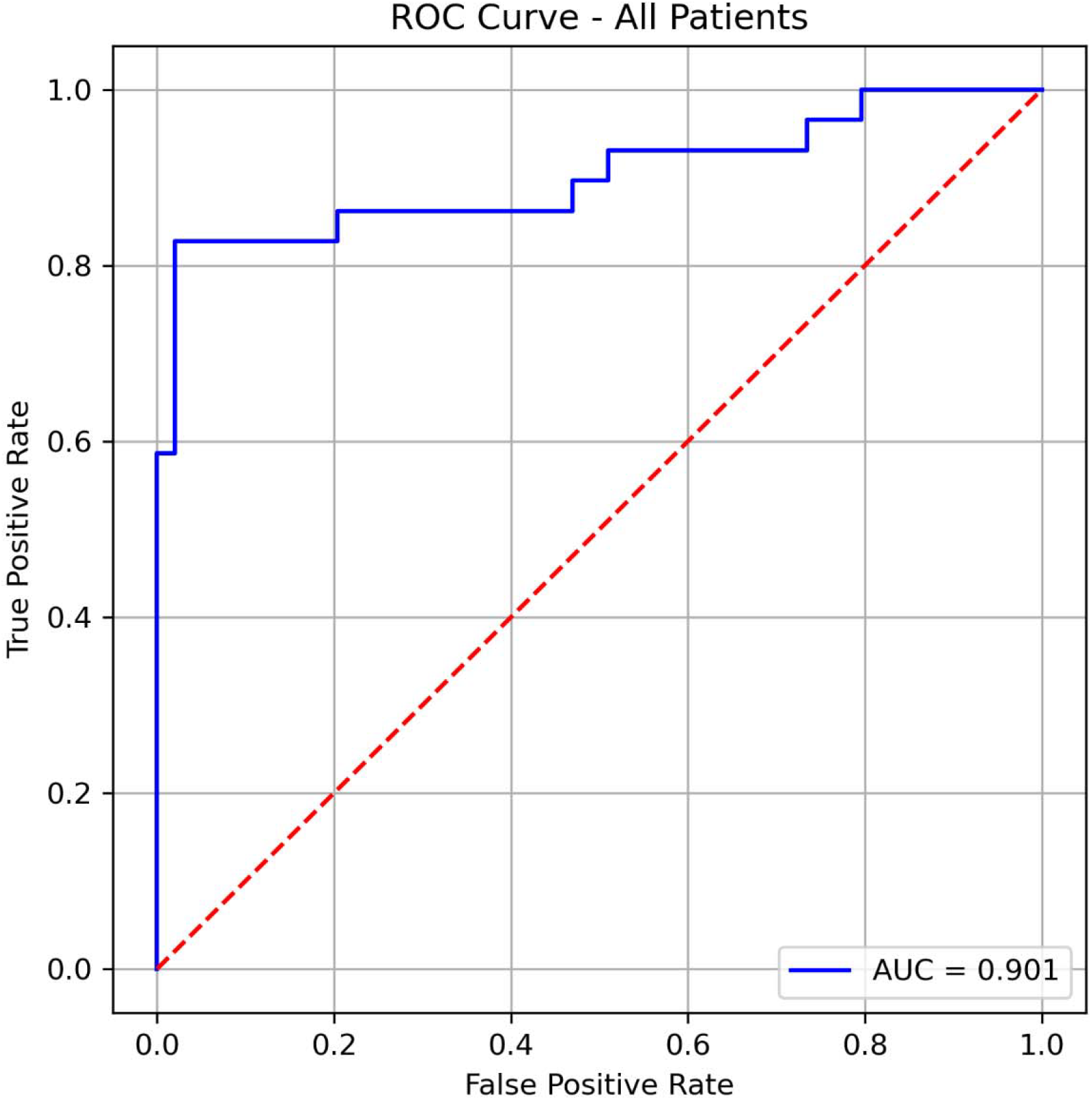
Receiver operating characteristic (ROC) curve of correctly labeling non-seizure freedom for a l 78 patients from three institutions. Each point on the ROC curve is calculated at a different threshold of the probability of the label. The area under the ROC curve (AUC) is 90.1%.

**Figure 3:**
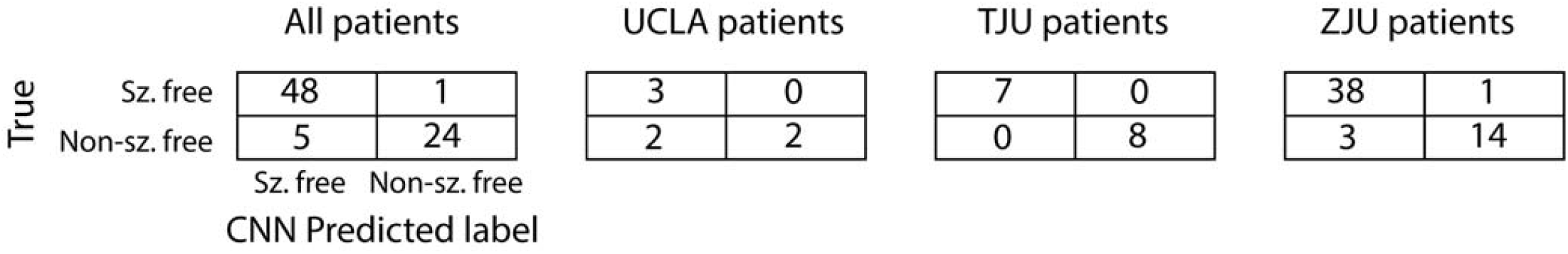
Confusion matrices comparing each patient’s actual label (seizure free [Sz. free] vs non-seizure free [Non-sz free]) compared to the label predicted by the three-branch convolutional neural network at the maximum of Youden’s J of the receiver operating characteristic (ROC) curve. Shown left to right are the confusion matrices for all the study patients, the UCLA patients only, the TJU patients only, and the ZJU patients only.

## Discussion

This research shows a method for prospectively improving epilepsy surgery outcomes by using a three-branch convolutional neural network (CNN) to predict post-operative seizure freedom. The CNN model was trained exclusively on diverse HFO features that were extracted from iEEG data recorded during non-REM sleep. These features were further analyzed within their neuroanatomic context and in relation to the proposed surgical margins. The resulting model achieved a predictive accuracy of 92%. Unlike traditional machine learning models that require flattening features into a single vector, a multi-branch CNN architecture can concurrently process heterogeneous data sources. For HFO analysis, this allows the model to separately handle per-electrode features, statistical measures across contacts, and resection status, thereby preserving critical inter-feature relationships. Branch 1 captures detailed HFO feature information at the level of individual electrodes. Branch 2 summarizes HFO feature global trends across electrodes by combining statistical and aggregated measures. Branch 3 encodes resection status, providing surgical context. When concatenated, the representation from Branch 3 influences how features from Branches 1 and 2 are interpreted, allowing the model to adjust the weight given to local or global patterns depending on whether electrodes were resected. This interaction between branches enables the network to focus on clinically meaningful patterns, which explains its improved performance over single-branch or conventional models.

The robust predictive performance of our CNN in classifying seizure freedom stems from 37 HFO related features. The capacity of the model to effectively harness these inputs is corroborated by a parallel univariate analysis, which independently demonstrated meaningful differences in these same HFO features between resected and unresected contacts across patients who did and did not achieve seizure freedom. While this confirms the value of a subset of the individual features, a systematic ablation study of the CNN is required to elucidate the relative contribution of individual features and feature groups to the CNN’s accuracy. Elucidating these key pathological biomarkers offering potential for mechanistic insight into epileptogenesis and ictogenesis and for the identification of novel therapeutic targets. featurefeaturefeaturefeaturefeature

The enthusiasm for machine learning and artificial intelligence to improve the outcome of epilepsy surgery is increasing[41] The accuracy of the CNN developed in our study for labeling seizure freedom is unprecedented. Prior investigations utilizing ML for predicting the seizure outcome of epilepsy surgery have utilized primarily clinical features [42], [43] A recent study used a linear model of HFO features to achieve 82% accuracy in predicting seizure free outcome, but this model included various clinical data features as well [44]. Our model did not utilize these clinical features and outperformed this model. Notably, machine learning has been used to study HFOs in the past but in contrast to labeling seizure freedom these ML strategies have sought to identify more pathological HFOs enriched in the SOZ or surgical margins of patients rendered seizure free [45], [46], [47], [48].

A future innovation of this framework is the use of counterfactuals—or virtual amendments to the surgical plan. If the CNN predicts that the initially proposed surgery is likely to fail, this system can generate and test alternative surgical plans to identify one with a higher probability of success. A future clinical trial could validate this approach by comparing two patient groups: a control arm receiving the standard-of-care resection and an experimental arm where the standard plan may be amended based on HFO metrics and counterfactual analysis. In the experimental arm, both the patient and the neurosurgeon would need to provide informed consent for the revised surgical plan[49], [50].

## Data Availability

All data produced in the present study are available upon reasonable request to the authors

