## Supplementary Material for "Multi-branch convolutional neural network using intracranial EEG high frequency oscillation features for predicting post-surgical seizure outcomes"

Supplementary Table 1: Univariate analysis of HFO features in the cohort of 49 patients rendered seizure free by surgery, p-value corrected with Holms-Bonferroni method. Abbreviations: RonO: ripple on oscillation; RonS: ripple on spike; fRonO: fast ripple on oscillation; fRonS: fast ripple on spike; RonO_rate_prior_spike: rate of RonO preceding epileptiform spikes <300 ms; fRonO_rate_prior_spike: rate of fRonO preceding epileptiform spikes <300 ms; UD/DU: ratio of HFO event occurring during at the Up-Down transition compared to Down-Up transition; RonS PAC zsore: Rayleigh Z measure of ripple coupling with epileptiform spike phase.

| HFO feature | Resected Mean | Resected Std. Err. | Unresected Mean | Unresected Std. Err | Corrected p-value | Cohen’s d effect size |
| --- | --- | --- | --- | --- | --- | --- |
| RonO_power_pk_stdev | 0.292 | 0.010 | 0.221 | 0.006 | 1.49E-06 | 1.206 |
| RonS_power_pk_z=3 | 7.060 | 0.069 | 6.697 | 0.044 | 8.44E-04 | 0.902 |
| RonO_power_pk_z=3 | 6.496 | 0.053 | 6.229 | 0.028 | 8.45E-04 | 0.898 |
| RonS_freq_pk_z=3 | 137.981 | 1.940 | 125.182 | 2.168 | 9.86E-04 | 0.892 |
| sharp_spike_rate | 1.021 | 0.200 | 0.154 | 0.030 | 1.45E-03 | 0.865 |
| RonS_power_pk_stdev | 0.400 | 0.016 | 0.324 | 0.011 | 5.69E-03 | 0.793 |
| fRonS_freq_pk_z=3 | 363.682 | 8.111 | 318.747 | 8.906 | 1.02E-02 | 0.779 |
| RonS_rate | 2.801 | 0.614 | 0.494 | 0.117 | 1.11E-02 | 0.746 |
| fRonO_power_pk_stdev | 0.256 | 0.008 | 0.217 | 0.008 | 3.02E-02 | 0.687 |
| RonO_rate_prior_spike | 0.454 | 0.121 | 0.065 | 0.014 | 5.36E-02 | 0.645 |
| fRonS_rate | 0.830 | 0.237 | 0.084 | 0.033 | 6.38E-02 | 0.631 |
| RonS_freq_pk_stdev | 14.038 | 0.577 | 11.469 | 0.625 | 8.49E-02 | 0.613 |
| fRonS_power_pk_z=3 | 5.921 | 0.063 | 5.673 | 0.060 | 1.34E-01 | 0.595 |
| RonO_fRonO_coincidence_rate | 0.986 | 0.258 | 0.249 | 0.050 | 1.48E-01 | 0.566 |
| fRonO_rate | 1.562 | 0.393 | 0.473 | 0.099 | 1.96E-01 | 0.542 |
| RonO_power_pk_mean | 5.824 | 0.026 | 5.750 | 0.013 | 2.84E-01 | 0.512 |
| fRonO_rate_prior_spike | 0.084 | 0.031 | 0.009 | 0.005 | 3.76E-01 | 0.487 |
| fRonO_power_pk_z=3 | 5.634 | 0.046 | 5.505 | 0.028 | 3.81E-01 | 0.484 |
| RonS_PAC_zscore | 47.123 | 11.147 | 19.896 | 5.514 | 5.70E-01 | 0.447 |
| RonO_freq_pk_z=3 | 145.019 | 2.198 | 139.383 | 1.553 | 6.99E-01 | 0.423 |
| RonS_power_pk_mean | 6.346 | 0.044 | 6.234 | 0.034 | 8.05E-01 | 0.408 |
| Delta UD/DU FastRipples | 1.538 | 0.132 | 1.224 | 0.084 | 7.63E-01 | 0.407 |
| RonO_rate | 16.221 | 2.171 | 11.615 | 0.893 | 7.89E-01 | 0.396 |
| RonO_freq_pk_stdev | 14.747 | 0.683 | 13.184 | 0.438 | 7.98E-01 | 0.389 |
| fRonS_freq_pk_stdev | 45.533 | 2.233 | 39.561 | 2.757 | 1.00E+00 | 0.373 |
| RonS_freq_pk_mean | 105.912 | 0.760 | 104.329 | 0.591 | 1.00E+00 | 0.335 |
| fRonO_freq_pk_z=3 | 347.184 | 7.867 | 332.329 | 6.305 | 1.00E+00 | 0.300 |
| fRonS_freq_pk_mean | 284.108 | 4.753 | 273.607 | 5.632 | 1.00E+00 | 0.298 |
| fRonS_power_pk_stdev | 0.257 | 0.016 | 0.228 | 0.016 | 1.00E+00 | 0.276 |
| Delta UD/DU Ripples | 1.489 | 0.104 | 1.329 | 0.055 | 1.00E+00 | 0.274 |
| fRonS_power_pk_mean | 5.501 | 0.034 | 5.434 | 0.044 | 1.00E+00 | 0.249 |
| fRonO_power_pk_mean | 5.244 | 0.029 | 5.205 | 0.015 | 1.00E+00 | 0.242 |
| Slow UD/DU FastRipples | 1.297 | 0.144 | 1.130 | 0.068 | 1.00E+00 | 0.216 |
| fRonO_freq_pk_stdev | 39.837 | 1.920 | 38.166 | 1.604 | 1.00E+00 | 0.136 |
| fRonO_freq_pk_mean | 272.602 | 4.675 | 269.048 | 3.200 | 1.00E+00 | 0.128 |
| Slow UD/DU Ripples | 1.645 | 0.126 | 1.706 | 0.067 | 1.00E+00 | -0.086 |
| RonO_freq_pk_mean | 106.705 | 0.999 | 106.748 | 0.643 | 9.71E-01 | -0.007 |

Supplementary Table 2: Univariate analysis of HFO features in the cohort of 29 patients not rendered seizure free by surgery, p-value corrected with Holms-Bonferroni method. Abbreviations: RonO: ripple on oscillation; RonS: ripple on spike; fRonO: fast ripple on oscillation; fRonS: fast ripple on spike; RonO_rate_prior_spike: rate of RonO preceding epileptiform spikes <300 ms; fRonO_rate_prior_spike: rate of fRonO preceding epileptiform spikes <300 ms; UD/DU: ratio of HFO event occurring during at the Up-Down transition compared to Down-Up transition; RonS PAC zsore: Rayleigh Z measure of ripple coupling with epileptiform spike phase.

| HFO feature | Resected Mean | Resected Std. Err. | Unresected Mean | Unresected Std. Err | Corrected p-value | Cohen’s d effect size |
| --- | --- | --- | --- | --- | --- | --- |
| RonO_power_pk_stdev | 6.552 | 0.071 | 6.193 | 0.026 | 3.22E-04 | 1.330 |
| RonS_power_pk_z=3 | 0.283 | 0.014 | 0.217 | 0.005 | 1.02E-03 | 1.238 |
| RonO_power_pk_z=3 | 0.291 | 0.048 | 0.086 | 0.016 | 3.37E-03 | 1.140 |
| RonS_freq_pk_z=3 | 7.173 | 0.092 | 6.732 | 0.067 | 7.67E-03 | 1.096 |
| sharp_spike_rate | 2.351 | 0.479 | 0.531 | 0.095 | 8.47E-03 | 1.058 |
| RonS_power_pk_stdev | 1.038 | 0.227 | 0.193 | 0.038 | 9.61E-03 | 1.045 |
| fRonS_freq_pk_z=3 | 0.426 | 0.021 | 0.331 | 0.016 | 1.67E-02 | 1.016 |
| RonS_rate | 5.856 | 0.029 | 5.742 | 0.013 | 1.58E-02 | 1.000 |
| fRonO_power_pk_stdev | 6.485 | 0.064 | 6.254 | 0.046 | 1.19E-01 | 0.829 |
| RonO_rate_prior_spike | 0.262 | 0.014 | 0.215 | 0.008 | 1.33E-01 | 0.825 |
| fRonS_rate | 44.729 | 8.134 | 22.606 | 5.881 | 7.73E-01 | 0.622 |
| RonS_freq_pk_stdev | 0.040 | 0.014 | 0.010 | 0.005 | 7.83E-01 | 0.602 |
| fRonS_power_pk_z=3 | 1.557 | 0.212 | 1.167 | 0.104 | 1.00E+00 | 0.492 |
| RonO_fRonO_coincidence_rate | 0.680 | 0.308 | 0.147 | 0.060 | 1.00E+00 | 0.483 |
| fRonO_rate | 0.683 | 0.250 | 0.297 | 0.057 | 1.00E+00 | 0.429 |
| RonO_power_pk_mean | 2.062 | 0.695 | 1.166 | 0.116 | 1.00E+00 | 0.397 |
| fRonO_rate_prior_spike | 17.233 | 2.232 | 13.647 | 1.262 | 1.00E+00 | 0.388 |
| fRonO_power_pk_z=3 | 133.409 | 2.654 | 128.490 | 2.400 | 1.00E+00 | 0.379 |
| RonS_PAC_zscore | 104.717 | 1.417 | 106.887 | 0.792 | 1.00E+00 | -0.371 |
| RonO_freq_pk_z=3 | 1.090 | 0.410 | 0.557 | 0.100 | 1.00E+00 | 0.358 |
| RonS_power_pk_mean | 0.292 | 0.028 | 0.258 | 0.019 | 1.00E+00 | 0.301 |
| Delta UD/DU FastRipples | 104.831 | 0.732 | 105.848 | 0.769 | 1.00E+00 | -0.261 |
| RonO_rate | 264.789 | 5.390 | 271.813 | 6.325 | 1.00E+00 | -0.226 |
| RonO_freq_pk_stdev | 1.656 | 0.241 | 1.472 | 0.078 | 1.00E+00 | 0.205 |
| fRonS_freq_pk_stdev | 12.765 | 0.650 | 12.055 | 0.757 | 1.00E+00 | 0.192 |
| RonS_freq_pk_mean | 12.442 | 0.640 | 12.969 | 0.428 | 1.00E+00 | -0.188 |
| fRonO_freq_pk_z=3 | 6.024 | 0.094 | 5.950 | 0.071 | 1.00E+00 | 0.187 |
| fRonS_freq_pk_mean | 320.175 | 10.958 | 328.148 | 9.244 | 1.00E+00 | -0.151 |
| fRonS_power_pk_stdev | 352.422 | 13.519 | 344.412 | 10.769 | 1.00E+00 | 0.137 |
| Delta UD/DU Ripples | 5.535 | 0.084 | 5.503 | 0.033 | 1.00E+00 | 0.099 |
| fRonS_power_pk_mean | 5.202 | 0.054 | 5.222 | 0.022 | 1.00E+00 | -0.097 |
| fRonO_power_pk_mean | 42.919 | 4.431 | 41.886 | 3.051 | 1.00E+00 | 0.058 |
| Slow UD/DU FastRipples | 34.617 | 2.775 | 35.379 | 2.479 | 1.00E+00 | -0.057 |
| fRonO_freq_pk_stdev | 1.730 | 0.212 | 1.686 | 0.120 | 1.00E+00 | 0.050 |
| fRonO_freq_pk_mean | 278.305 | 6.968 | 279.860 | 7.288 | 1.00E+00 | -0.044 |
| Slow UD/DU Ripples | 138.358 | 2.487 | 138.523 | 1.416 | 1.00E+00 | -0.016 |
| RonO_freq_pk_mean | 5.575 | 0.048 | 5.576 | 0.040 | 9.81E-01 | -0.007 |
